# The Impact of Electrode Placement and Electrical Conductivity Uncertainties on Temporal Interference Stimulation

**DOI:** 10.64898/2026.01.27.26344919

**Authors:** Sicheng An, Luca Di Rienzo, Lorenzo Codecasa, Thomas R. Knösche, Axel Thielscher, Konstantin Weise

## Abstract

Temporal interference stimulation (TIS) promises deeper and more selective neuromodulation, yet predictions remain sensitive to uncertainties in electrode setup and head modeling. We investigate the impact of coregistration error (CE) of the volume conductor and the head, electrode placement uncertainty (EP), and tissue conductivity uncertainty (CU) on the electric field generated by TIS. The stochastic model aggregates CE, EP, and CU into nineteen random variables and is evaluated for a deep target in the left hippocampus and a superficial target in the motor cortex. The uncertainty and sensitivity analysis of the maximal modulation envelope of the electric field is based on an adaptive polynomial chaos expansion (PCE). Spatial statistics show that the mean of the electric field remains focalized over the regions of interest (ROIs), whereas the standard deviation is concentrated in targeted regions, indicating that uncertainty perturbs the electric field magnitude more than its focality. Variance decomposition reveals a clear hierarchy: CU is the main contributor to field variability, EP has a modest influence, and CE is essentially negligible within the considered ranges. Probability-density estimates of the mean strength inside and outside ROIs demonstrate separated distributions, confirming strong dose selectivity for both deep and superficial targets, while the relative difference measure (RDM) confirms that the spatial pattern of the TI field stays close to the reference configuration, indicating that uncertainty rescales the field rather than relocating its focal point. Generalization across four individualized head models spanning a range of head sizes confirms that the variance hierarchy CU > EP > CE and the dominance of a single electrode within EP are preserved across anatomies. Overall, within realistic modeling and setup uncertainties, TIS targeting appears robust when using state-of-the-art MRI-based electrode localization. The analysis identifies insufficient knowledge of tissue conductivities as the primary limitation for further improving the reliability of electric field predictions in TIS.

## I. Introduction

**T**emporalinterference stimulation (TIS) has emerged as a promising non-invasive brain stimulation technique, capable of achieving deeper and more selective neuromodulation than conventional approaches [1]–[3]. TIS usually uses two pairs of electrodes delivering high-frequency currents with slightly different frequencies [4]. While the high-frequency currents themselves weakly affect brain activity, their superposition produces a low-frequency amplitude modulation envelope that can influence neuronal activity [5], [6]. Since the envelope is widely regarded as the primary driver of neural modulation in TIS, assessing its spatial distribution is essential for predicting both the focality and stimulation strength. In what follows, we refer to the maximal modulation envelope distribution as the temporal interference (TI) field.

In practice, several factors perturb the TI field and thereby introduce uncertainty into electric field predictions [7]–[9]. In this work we focus on three broad and practically relevant categories that occur in state-of-the-art electrode position optimization and placement based on an individual MRI and neuronavigation: (i) coregistration error (CE), defined as translation or rotation of the head relative to the volume conductor model, e.g. caused by inaccuracies of registration procedure of the head in neuronavigation devices and instability of the head reference marker [10], [11]; (ii) electrode placement uncertainty (EP), defined as variability in electrode locations on the scalp [12]–[15]; and (iii) tissue conductivity uncertainty (CU), defined as interindividual and intraindividual variability in the electrical properties of head tissues [16]–[18]. Although optimized electrode placement and configuration procedures exist [19]–[22], they typically neglect these uncertainties, which limits the generalizability of predicted effects.

As various uncertainty sources need to be considered simultaneously, we are challenged by having to analyze a high-dimensional model. Traditional uncertainty analysis methods, such as Monte Carlo simulation, become prohibitive due to computationally expensive finite element solutions for electric field prediction. To address this challenge, we adopt an adaptive polynomial chaos expansion [23], [24] to construct a surrogate model that captures the dependence of the TI field and related quantities of interest (QoIs) on the uncertain inputs. This surrogate model enables efficient statistical property estimation of QoIs and variance-based global sensitivity analysis, i.e., Sobol indices. While polynomial chaos expansion (PCE) has been successfully applied in previous studies to other established stimulation modalities [18], [25], the application to TIS presents fundamentally distinct scientific and methodological challenges.

In this paper, we propose a comprehensive framework for uncertainty and sensitivity analysis tailored to TIS. To the best of our knowledge, this is the first study to simultaneously develop and integrate stochastic models for CU, CE, and EP. To accurately simulate the geometric complexities of CE and EP on human head models, we adapt an ellipsoid coordinate system to successfully map these spatial shifts. Spatial targeting is characterized using region-wise and element-wise statistics of the electric field. We use the framework for two representative targets (left hippocampus and left motor cortex) and report spatial statistics of the electric field, decompose variance across uncertainty categories and individual parameters, and estimate the probability distributions of the mean stimulation strength inside and outside ROIs, and quantify the spatial stability of the TI field via the relative difference measure (RDM). To assess whether the main findings depend on a single anatomy, we further extend the analysis to three additional individualized head models, spanning the variability of head size.

The remainder of the paper is organized as follows. Section II details the modeling of uncertain sources, the calculation of the electric field, the procedures of uncertainty and sensitivity analysis, and the definitions of the metrics for evaluating TIS effects. Section III presents the results, including spatial statistics, variance decompositions by category and parameter, probability density estimates of the stimulation metrics and of the spatial stability measure, and a generalization analysis across four individualized head models. Section IV discusses the findings, provides practical recommendations for the TIS development, and outlines limitations and directions for future work. Section V concludes the paper.

## II. Methods

### A. Stochastic Modeling

To describe CE, i.e., the rigid-body misalignment of the volume conductor with respect to head, in a way that is smooth and anatomically plausible, the skin surface is approximated by a triaxial ellipsoid fitted in the least-squares sense to the nodes of the valid skin surface, as shown in Fig. 1(a)–(b) [19]. In practice, a given CE realization corresponds to a specific perturbation of this ellipsoid, representing a plausible shift and reorientation of the entire skin surface rather than the independent motion of individual nodes. This uncertainty is parameterized by six variables: three translation distances of the ellipsoid center (Δ _*x*_, Δ _*y*_, Δ _*z*_) and three rotation angles around the axes (*θ*_*x*_, *θ*_*y*_, *θ*_*z*_). Examples of translation and rotation are illustrated in Fig. 1(c)–(d). For clarity, the magnitudes shown are intentionally exaggerated and do not represent the actual uncertainty ranges considered in our analysis.

**Fig. 1.**
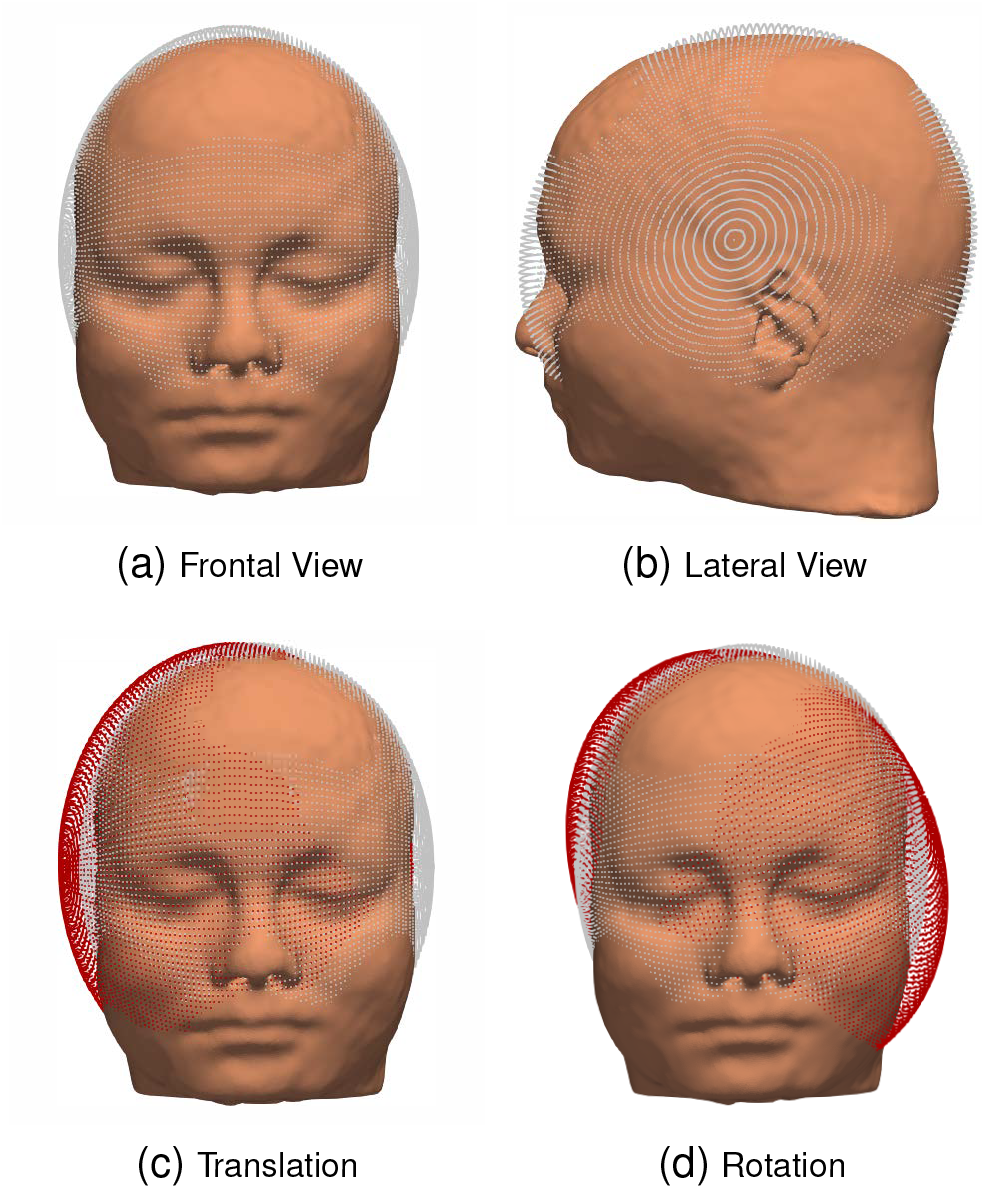
Ellipsoid-based representation of coregistration error. A triaxial ellipsoid is fitted to the skin surface and then (c) translated and (d) rotated to parameterize uncertainties. For visual clarity, the translation and rotation magnitudes are exaggerated compared to the actual uncertainty ranges in Table I.

EP represents the uncertainty in electrode placement on the scalp, arising from electrode drift and subtle manual positioning variability during preparation. It is modeled on the transformed ellipsoid corresponding to the current CE sample. Each initial electrode location on the skin surface is first projected onto the ellipsoid along the outward normal of the skin surface [19]. For electrode *k* with *k* = 1, …, 4, the perturbation is described by a geodesic displacement distance *d*_*k*_ and a direction *ϕ*_*k*_ on the ellipsoid surface. In practice, a given EP realization therefore corresponds to a plausible tangential misplacement of each electrode along the scalp, relative to its nominal target position, rather than a change in its distance to the skin. The perturbed ellipsoid position is obtained by solving the geodesic problem with initial point at the projected nominal location and initial tangent set by *ϕ*_*k*_ [19], [26], as illustrated in Fig. 2. For visual clarity, the electrode displacements shown in the figure are intentionally larger than the typical uncertainty levels considered. The perturbed electrode is then mapped back to the skin by reverse projection, and the updated position is used in the electric field calculation.

**Fig. 2.**
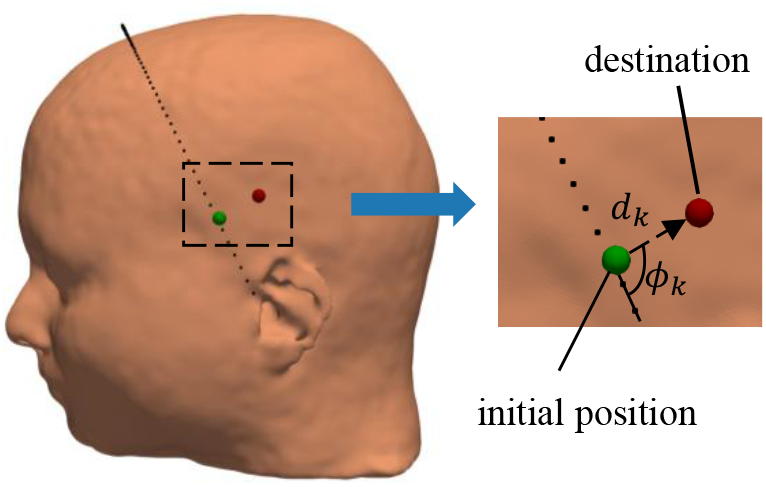
Electrode placement uncertainty modeled as a geodesic displacement on the fitted ellipsoid. From an initial location, each electrode is moved by distance ***d***_***k***_ along direction ***ϕ***_***k***_, yielding a destination obtained by solving the ellipsoidal geodesic problem [19]. For visual clarity, the electrode placement uncertainty is exaggerated relative to typical values.

The anatomical head model used in this work is the Ernie model, generated with the CHARM pipeline [27] as implemented in SimNIBS [28]. To better resolve variability in electrode placement on the scalp, the skin surface is remeshed at a higher resolution before finite-element discretization. The resulting head model consists of approximately 6.71× 10^6^ tetrahedral elements and 1.01 × 10^6^ vertices, and a single FEM calculation of the TI field on the midlayer gray matter surface on this refined head model requires about 688 s of computation time. The head model is shown in Fig. 3.

**Fig. 3.**
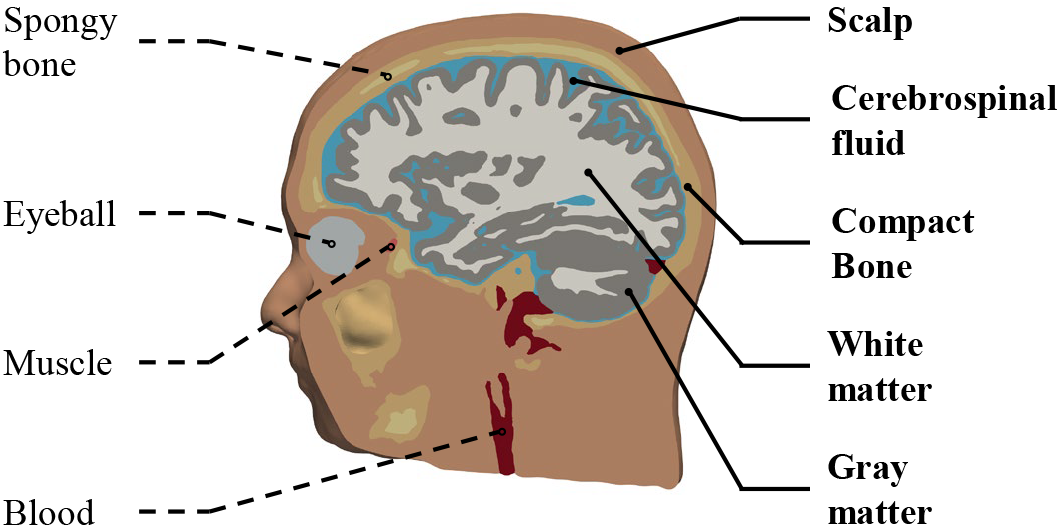
Ernie head model used in this paper. The bold tissues were included in the uncertainty analysis as they were identified to be dominant in [18].

CU represents the uncertainty in tissue conductivity values within the head model, arising from inter-subject variability and measurement uncertainty in tissue electrical properties. Based on [18], the conductivities of white matter (WM), gray matter (GM), cerebrospinal fluid (CSF), compact bone (CB), and scalp (S) are modeled explicitly as uncertain parameters, because these tissues account for the majority of the variability in the electric field on the midlayer GM surface. The conductivities of the remaining tissues are fixed to established values reported in [29]–[31].

In total, we model 19 mutually independent random inputs describing CE, EP, and tissue conductivity: six variables for CE, three translations of the ellipsoid center (Δ _*x*_, Δ _*y*_, Δ _*z*_) and three rotations (*θ*_*x*_, *θ*_*y*_, *θ*_*z*_); eight variables for EP, parameterized per electrode by the geodesic displacement distance and direction (*d*_*k*_, *ϕ*_*k*_), *k* = 1, …, 4; and five variables for tissue conductivities.

The CE and EP parameters are modeled as independent uniform random variables over the bounds listed in Table I. The translation and rotation ranges for CE are chosen to reflect typical coregistration inaccuracies reported for head-MRI registration and instability of the head tracker, while excluding clearly unrealistic misalignment [10], [11]. The boundaries of the EP also encompass the placement variability observed in TES stimulation under the latest monitoring methods [12]– [14], representing reasonable displacement of electrodes on the scalp surface rather than severe misplacement.

**TABLE I.** Summary of uncertain Parameters for Stochastic Modeling.

| Parameter | Distribution | Min | Max | Reference |
| --- | --- | --- | --- | --- |
| $\Delta x, \Delta y, \Delta z$ | Uniform | −2 mm | 2 mm | [10], [11] |
| $\theta_x, \theta_y, \theta_z$ | Uniform | −1° | 1° | [10], [11] |
| $d_1, d_2, d_3, d_4$ | Uniform | 0 mm | 4 mm | [12]–[14] |
| $\phi_1, \phi_2, \phi_3, \phi_4$ | Uniform | 0° | 360° | [12]–[14] |
| $\sigma_{WM}$ | Beta(3, 3) | 0.1 S/m | 0.4 S/m | [32]–[34] |
| $\sigma_{GM}$ | Beta(3, 3) | 0.1 S/m | 0.6 S/m | [32], [35]–[37] |
| $\sigma_{CSF}$ | Beta(3, 3) | 1.2 S/m | 1.8 S/m | [30], [38] |
| $\sigma_{CB}$ | Beta(3, 3) | 0.003 S/m | 0.014 S/m | [39], [40] |
| $\sigma_S$ | Beta(3, 3) | 0.2 S/m | 0.5 S/m | [30], [41], [42] |

For tissue properties, each conductivity *σ*_*i*_ (WM, GM, CSF, CB, S) is assigned an independent scaled Beta distribution 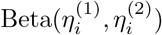 on (*a*_*i*_, *b*_*i*_) [18]. The corresponding probability density function (PDF) is given by

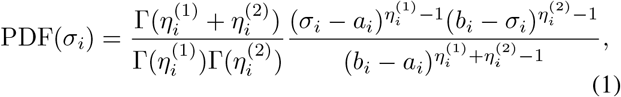

where (*a*_*i*_, *b*_*i*_) denote the conductivity bounds, 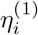and 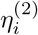 are the shape parameters, and Γ(·) denotes the Gamma function. Specifically, we set the parameters to 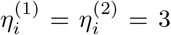. The conductivity distributions provide symmetric, bell-shaped priors centered on the nominal conductivity values while ensuring that all samples remain within physiologically plausible limits. The choice follows the uncertainty modeling adopted in previous studies on TES and TMS [18], enabling direct comparability with earlier work. The intervals (*a*_*i*_, *b*_*i*_) are selected to robustly cover the values reported in the experimental literature [30]–[42]. We only consider studies that measured relatively fresh or in vivo tissue (preferably human) at low frequencies (0–100 kHz) and near body temperature. All random inputs, their distribution functions, and the corresponding bounds are summarized in Table I.

### B. Electric Field Calculation

We calculate the electric field generated by each pair of electrodes by means of the first-order tetrahedral finite element method combined with the super-convergent patch recovery (SPR) implemented in SimNIBS [43]. In contrast to the conventional FEM pipeline for TIS that embeds electrodes as volumetric domains, we model electrodes as surface current sources by imposing von Neumann boundary conditions at the relevant skin nodes. This modeling choice avoids repeated and costly mesh modification steps associated with volumetric electrode insertion [19].

The use of Neumann boundary conditions requires specifying the node currents at the skin interface corresponding to the electrode area. The relevant nodes are identified as those located on the outer surface of the skin mesh within the projected electrode footprint. For each of these nodes, the associated surface area is computed from the surrounding surface elements, which allows determining the corresponding current contribution based on the local current density.

However, applying a constant current density does not correctly represent the underlying electric field problem for TI. In practice, the injected current is limited or controlled by the stimulator, while each conductive electrode behaves approximately as an equipotential surface. While modeling constant electrode potentials can be achieved by applying Dirichlet boundary conditions to the corresponding nodes, this requires updates of the stiffness matrix and repetition of the solver preparation steps whenever electrode positions change, which substantially increases computational cost.

To avoid repeatedly imposing Dirichlet boundary conditions, we instead solve a secondary optimization problem that iteratively adjusts the node currents associated with the electrode area such that the condition of constant electric potential across each electrode is satisfied. Details of this current-tuning approach are described in Supplemental Material S1 of [19]. Although this procedure requires repeated FEM simulations, it remains significantly more efficient than rebuilding and preconditioning the system matrix for each electrode configuration.

Let **E**_1_(**r**) and **E**_2_(**r**) denote the electric fields at the location **r** generated by the first and second electrode pair, respectively. The maximal amplitude of the modulation envelope of the superimposed field at the location **r** is then computed as [4]:

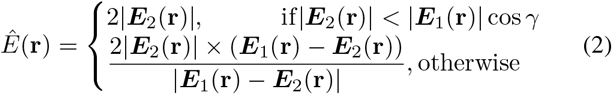

where *γ* is the angle between **E**_1_(**r**) and **E**_2_(**r**) . Throughout this work, we take *Ê*(**r**) as the primary quantity of interest and, for brevity, refer to it simply as the TI field.

### C. Uncertainty and Sensitivity Analysis

The PCE provides an efficient surrogate modeling framework to represent the dependence between the random variables ***ξ*** and the QoIs **q** = [*q*_1_, · · ·, 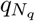], where *N*_*q*_ is the number of QoIs. Specifically, each QoI *q*_*i*_(***ξ***) is approximated in terms of an orthonormal polynomial basis as:

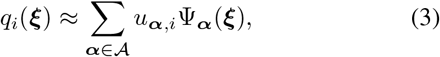

where 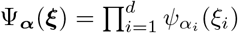 is a multivariate basis function constructed from univariate orthogonal polynomials 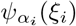, and ***α*** is a multi-index denoting the polynomial degrees. *A* denotes the finite set of multi-indices corresponding to the finite polynomial basis set.

In this work, we employ an adaptive PCE strategy, which iteratively enriches the polynomial basis set *A* until a target accuracy is reached. The general procedure can be summarized as follows:

- **Polynomial family selection:** identify the orthonormal polynomial families associated with the input distributions, following [23].
- **Initialization:** construct an initial polynomial basis, generate random samples of input variables, and calculate the corresponding QoIs.
- **Basis extension:** extend the active basis set using a basis-increment strategy.
- **Samples update:** add new samples of input variables and their corresponding QoIs to maintain the ratio between the number of samples and the number of basis terms.
- **Coefficient calculation and error estimation:** compute PCE coefficients and assess approximation error.
- **Stopping criteria:** terminate once the target accuracy is achieved or a maximum iteration limit is met; otherwise, continue extension.

The PCE coefficients are determined using a regression-based approach. Let 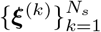 be the samples of the input uncertain parameters, ***Q*** be the matrix of QoI with entries 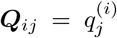 which denotes the *j*th QoI *q*_*j*_ of the *i*th input sample ***ξ***^(*i*)^, and **Ψ** be the matrix of basis function with entries 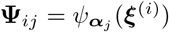. The PCE coefficients are estimated by

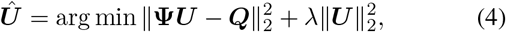

where *λ* denotes the regularization parameter, which is chosen among nine log-spaced values between 10^−5^ and 10^3^.

The basis extension follows the adaptive grid quadrature algorithm [44]. Multi-indices adjacent to the index with the largest variance contribution are added to the set. The contribution is evaluated by

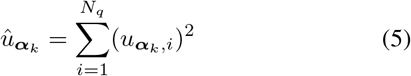

which is proportional to the fraction of variance explained by the corresponding polynomial basis function 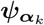 Collectively, the contribution vector is denoted as 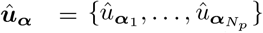, where *N*_*p*_ is the total number of polynomial terms. At each iteration, the index *α*_*k*_ ′ with the largest 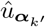 is selected, and all its forward neighbors that preserve downward closedness are added to the basis set. The resulting anisotropic extension algorithm is summarized in Algorithm 1. During the basis extension, we maintain two dynamically updated sets of multi–indices:

- the candidate set *S*_*cand*_ containing multi-indices that can still be expanded;
- the old set *S*_*old*_, containing multi-indices that have already been expanded.

The approximation error is estimated using an independent validation set *X*_*v*_ of *N*_*v*_ randomly drawn samples. The error

#### Algorithm 1 Anisotropic basis extension for adaptive PCE

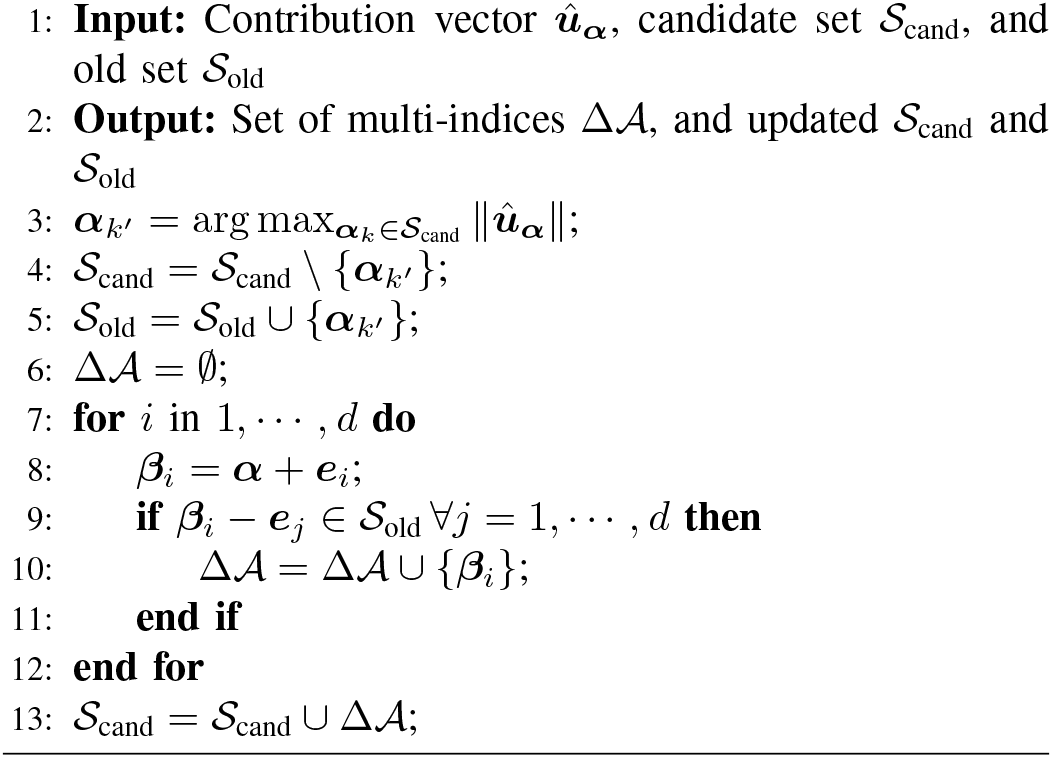

metric is defined as:

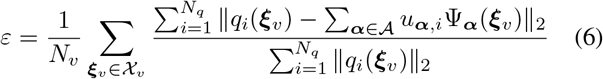

Once the PCE model is obtained, statistical quantities can be calculated directly from the PCE coefficients. For example, the mean of the QoI *q*_*i*_ is determined by:

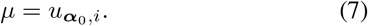

Similarly, the variance is determined by:

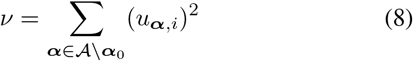

where ***α***_0_ = [0, 0, · · ·, 0]. To preserve measurement consistency in reporting, we quantify uncertainty using the standard deviation, defined elementwise as 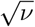. The relative standard deviation (RSD) with respect to the expectation is then given by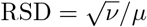.

Beyond statistical estimation, PCE also facilitates variance-based sensitivity analysis. The Sobol indices decompose the total variance into contributions from individual input parameters or their interactions [45], [46]. For each QoI *q*_*i*_, the group Sobol indices of a subset *G* of variables are given by:

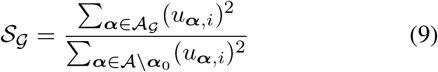

Where *A*_*G*_ ⊂ *A* consists of the indices with nonzero entries only for variables in *G*, covering all polynomial orders and all within-group interactions. The Sobol indices are normalized with respect to the total variance and consequently add up to one. In some cases, however, non-normalized indices are preferable, particularly when analyzing variances and means of different magnitudes, as encountered in field distributions at locations with either strong or negligible field intensities.

### D. Metrics for Evaluating TIS Effects

To complement the analysis of the electric field, we introduce quantitative metrics that capture the stimulation strength and spatial stability. Let Ω_*ROI*_ denote the target region, Ω_*mid*_ the midlayer GM, and Ω_*out*_ the part of the midlayer GM excluding the target region.

The stimulation strength is evaluated inside and outside the target region, for the uncertain input sample ***ξ***^(*k*)^, the mean stimulation strength within the target region is defined as

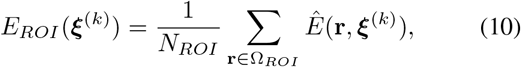

where *N*_*ROI*_ denotes the number of elements in the target region and *Ê*(**r, *ξ***^(*k*)^) denotes the TI field for ***ξ***^(*k*)^ at location **r** . Similarly, the mean stimulation strength outside the target region is defined as

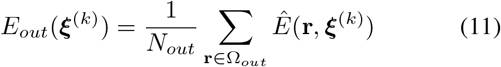

where *N*_*out*_ is the number of elements in the midlayer gray matter excluding the target region.

In addition to these magnitude-based metrics, we quantify the spatial stability of the TI field under uncertainty using the relative difference measure (RDM) [47], evaluated over the whole midlayer GM Ω_*mid*_. The midlayer GM was chosen because the present analysis focuses on the cortical neuron, which are situated in the GM. For the *k*th uncertain input sample ***ξ***^(*k*)^, the RDM is defined as

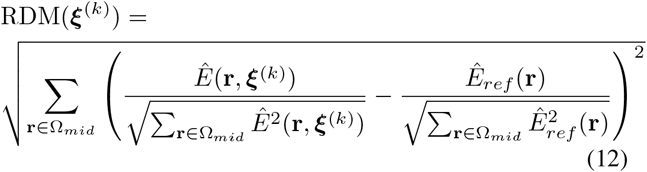

where *Êref* (**r**) is the TI field corresponding to the reference configuration, i.e., zero coregistration error, nominal electrode positions, and reference tissue conductivity values. Since both fields are normalized before comparison, the RDM isolates changes in the spatial field distribution from changes in the overall field magnitude. An RDM of zero indicates identical normalized spatial distributions, whereas larger values indicate increasing spatial distortion. Values in the range of 0–0.3 generally indicate that the normalized field patterns remain similar, while values approaching 1 indicate substantial distortion of the normalized field pattern.

## III. Results

### A. Overview

We use a leadfield-free optimization method [19] to determine the electrode positions targeting the left hippocampus and left motor cortex. Each pair comprised two circular electrodes with diameters 20 mm, and a peak current of *I*_max_ = 2 mA was applied for both targets. For the hippocampus case, electrode pairs were placed at (™ 75, 33, ™38) mm with (™75, 28, ™ 4) mm, and (−20, 113, 23) mm with (65, 84, 11) mm ; for the motor cortex case, electrode pairs were at (™49, 68, 60) mm with (49, ™21, 84) mm, and (™32, 7, 91) mm with (™50, ™11, 82) mm, as shown in Fig. 4.

**Fig 4.**
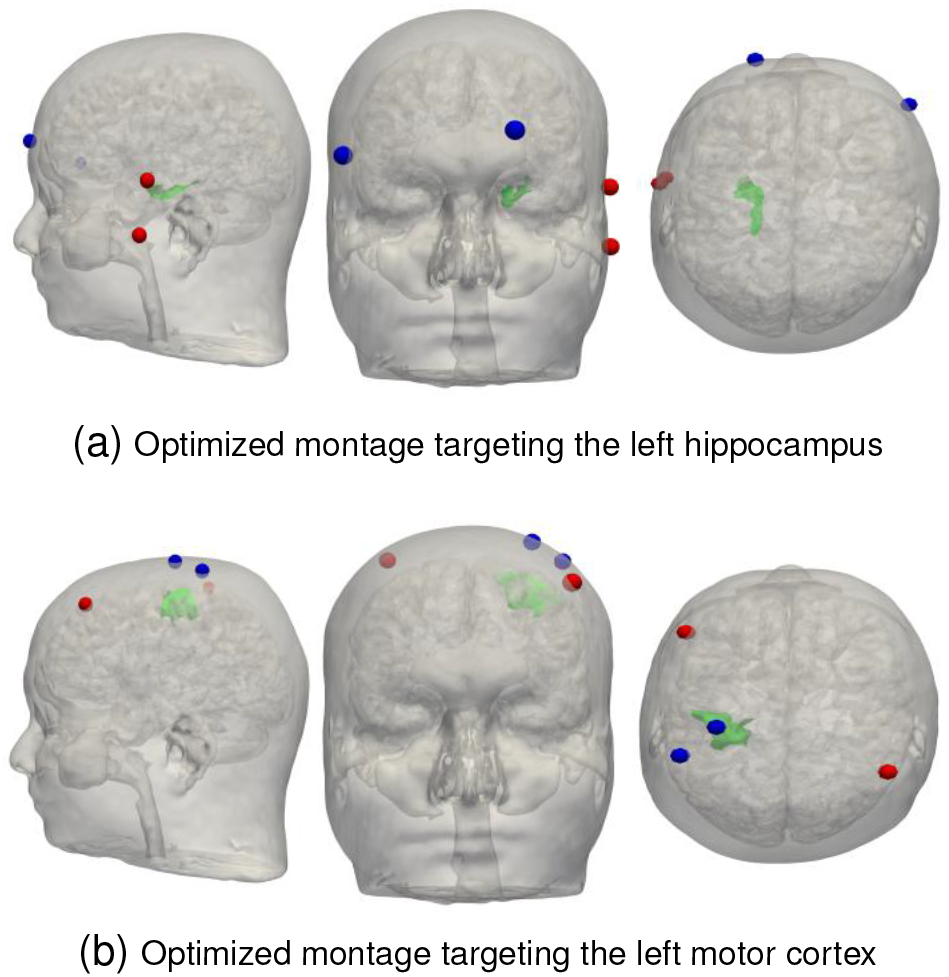
Optimized electrode montages for targeting left hippocampus and left motor cortex. The green region denotes the ROI; red and blue markers indicate the first and second electrode pairs, respectively.

Deterministic simulations showed that, for the left hippocampus case, the mean electric field magnitude in the ROI is 0.237 V/m versus 0.094 V/m outside the ROI. For the motor cortex case, the mean within-ROI strength was 0.316 V/m versus 0.089 V/m outside.

For the electric field within the ROIs, the adaptive PCE models converge after 2812 simulations for the hippocampus case and 2744 for the motor cortex case. For the electric field on the midlayer GM, the adaptive PCE models achieve validation errors *<* 10^−2^ for the hippocampus case with 2252 simulations, whereas for the motor cortex case, the validation error is 1.12 × 10^−2^ after 3000 simulations. The corresponding maximum polynomial and interaction orders are summarized in Table II.

**TABLE II.**
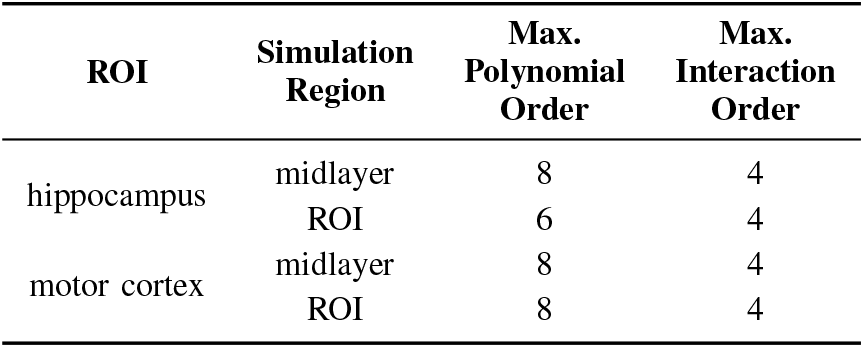
Maximum Polynomial Orders and Interaction Orders of the PcE Basis Constructed for the ti Field on the Midlayer gm and the roi Surface for Each Target. The Maximum polynomial order is defined as the maximum order of any individual polynomial, and the maximum interaction order is the maximum number of different variables involved in a polynomial basis.

### B. Spatial Statistics

The spatial statistics of the TI field strength in the middle cortical sheet and in the ROIs are shown in Fig. 5, where the mean represents the expected field magnitude, the standard deviation quantifies the absolute uncertainty at each location, and the relative standard deviation characterizes the uncertainty normalized by the local stimulation strength. The mean distribution is strongly focused within the ROIs and remains consistent across the different targets, indicating robust targeting performance. The standard deviation is likewise concentrated in regions of high intensity, showing that most of the absolute variability is confined to the stimulated areas. Although the relative standard deviation is larger in low-intensity off-target regions, this primarily reflects normalization by a small mean field: in absolute terms, these regions remain weakly stimulated and do not approach the field levels observed in the ROIs in any of the considered realizations.

**Fig 5.**
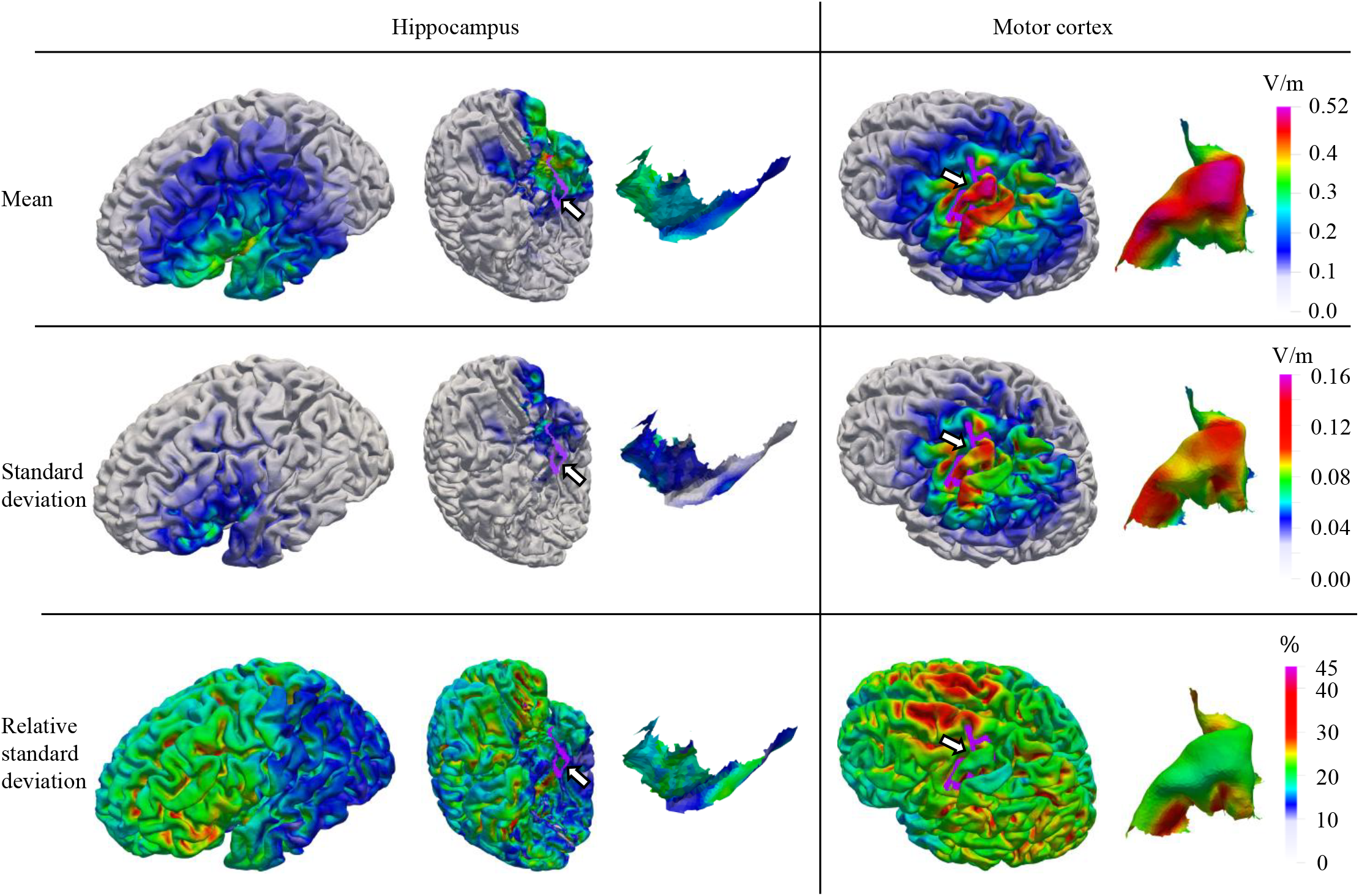
Mean, standard deviation, and relative standard deviation of the TI field on the middle GM and ROI surfaces. To compare the different considered targets, all mean maps share the same color scale, and all standard deviation maps share another. Rows correspond to statistical measures, and columns correspond to the stimulation target. The white arrows and purple circles indicate the ROI from which the cut-out surface is extracted.

### C. Sensitivity Analysis

The individual contributions of each uncertainty category (CE, EP, CU, and residual terms) are represented by Sobol indices. The element-wise contributions are shown in Fig. 6, and the aggregated contributions of each category are reported in the first column in Fig. 7. From Fig. 6, CU is seen to account for the majority of the field variance, with its influence extending over almost the entire midlayer GM surface and being particularly pronounced in regions of high intensity. In contrast, CE and EP not only have a smaller overall impact on the stimulation intensity but also exhibit more localized effects, primarily confined to the target area and its surrounding regions. The interaction between each group basically has no impact on the electric field variation. This is corroborated by the variance decomposition in Fig. 7, where CU contributes approximately 90% of the variance, EP contributes 7–9%, and CE and the residual terms each contribute only about 1%. These patterns are even more pronounced when stimulating the motor cortex, consistent with the higher field strengths and greater spatial sensitivity in cortical targets.

**Fig 6.**
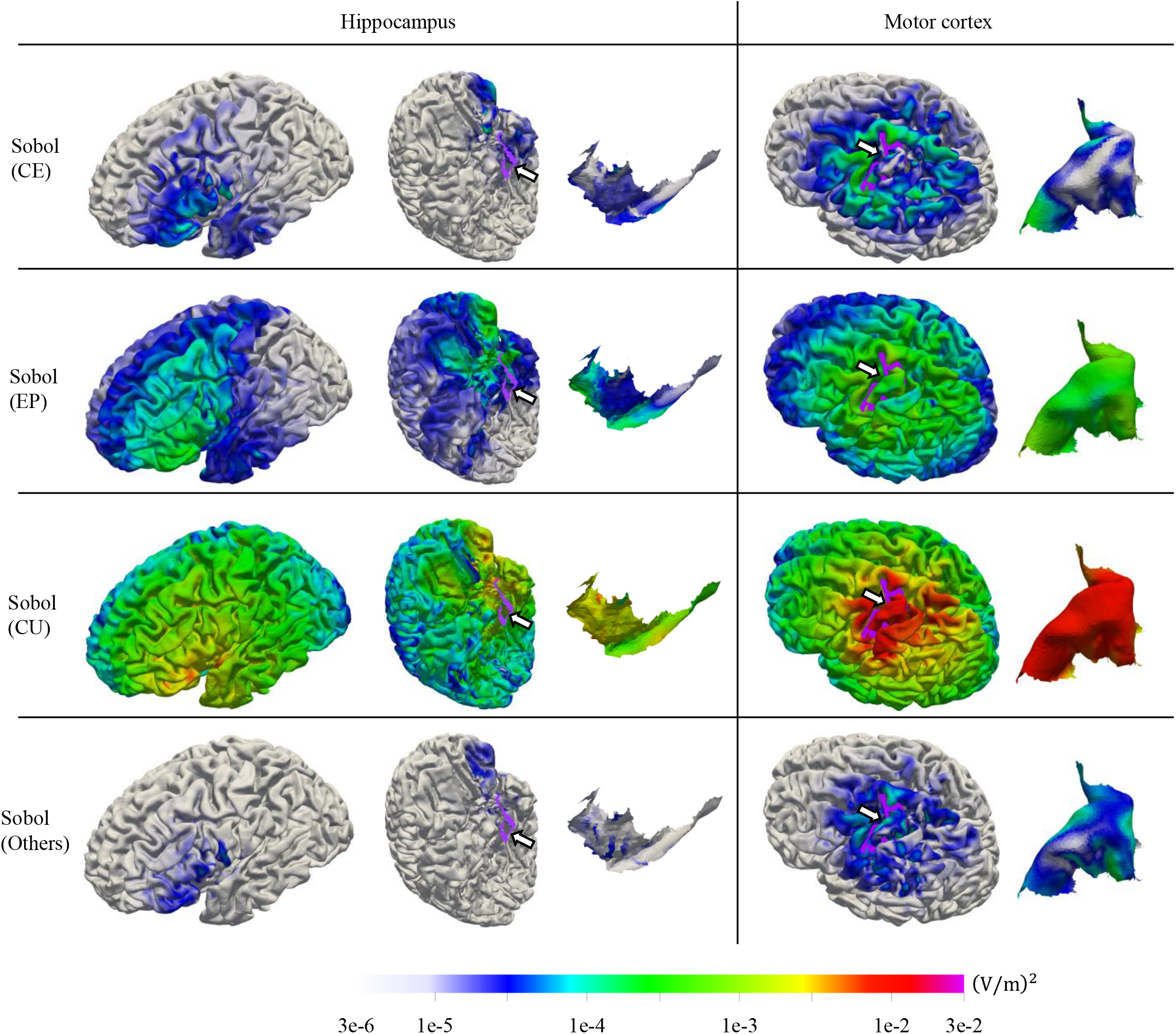
Sobol indices associated with each uncertainty category. All Sobol indices maps share the same color scale to provide comparability. We report unnormalized (partial variances) contributions rather than variance-normalized Sobol indices because the local field variance varies widely across space. The arrow and purple circle indicate the ROI from which the cut-out surface is extracted.

**Fig 7.**
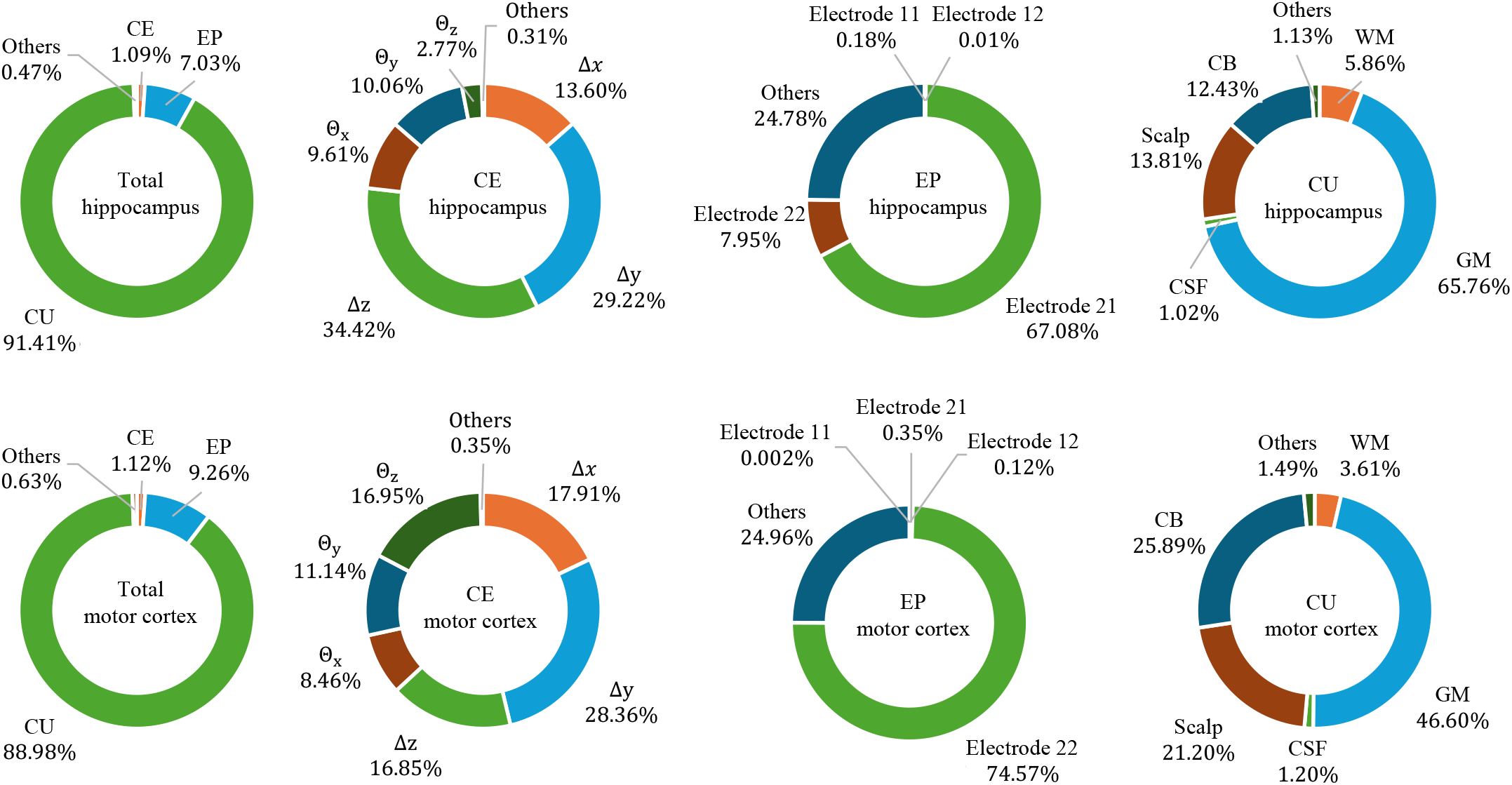
Relative contribution of uncertainty categories and individual parameters within each category. Electrode identifiers follow a two-digit code ***ij*** where the first digit ***i*** indexes the electrode pair and the second digit ***j*** indexes the electrode within that pair. The contributions are normalized to 100%

The contributions of individual parameters within each uncertainty group are also reported in Fig. 7. Within CE, translations contribute more strongly than rotations, and their interaction terms are minor. For the hippocampus case, translations along Δ*y* and Δ*z* dominate, and rotations about *θ*_*x*_ and *θ*_*y*_ are most influential. For the motor cortex, the three translation axes contribute comparably (with a slight dominance of Δ*y*), and the rotational contributions are similarly balanced, with a somewhat larger share for *θ*_*z*_. Within EP, where distance and direction are considered jointly, the electrode-level decomposition reveals strong asymmetries: in the hippocampus case, a single electrode (ID 21) accounts for 67.08% of the EP variance, and in the motor cortex case, a single electrode (ID 22) accounts for 74.57%, while the remaining electrodes contribute negligibly. Higher-order interactions between electrodes also play an important role, contributing 24.78% of the EP variance in the hippocampus case and 24.96% in the motor cortex case. Within CU, uncertainty in gray-matter conductivity contributes the most (65.76% for the hippocampus case and 46.60% for the motor cortex case), followed by the scalp (13.81% and 21.20%, respectively) and compact bone (12.43% and 25.89%, respectively). Cross-tissue interaction terms and CSF have negligible impact, each contributing only about 1%.

### D. Stimulation Magnitude and Spatial Stability

To complete the uncertainty analysis, we estimated the PDFs of the three metrics defined in Section II-D directly from the 4000 FEM samples used for PCE model construction and validation by means of kernel density estimation (KDE).

The distributions of *E*_*ROI*_ and *E*_*out*_ are shown in Fig. 8. For both targets, the distribution of *E*_*ROI*_, defined in (10), is shifted toward higher field strengths compared to *E*_*out*_, defined in (11), with *E*_*out*_ concentrated at lower values. Although a little overlap between the distributions remains, *E*_*ROI*_ is, on average, substantially larger than *E*_*out*_, indicating overall dose selectivity in favor of the ROI.

**Fig 8.**
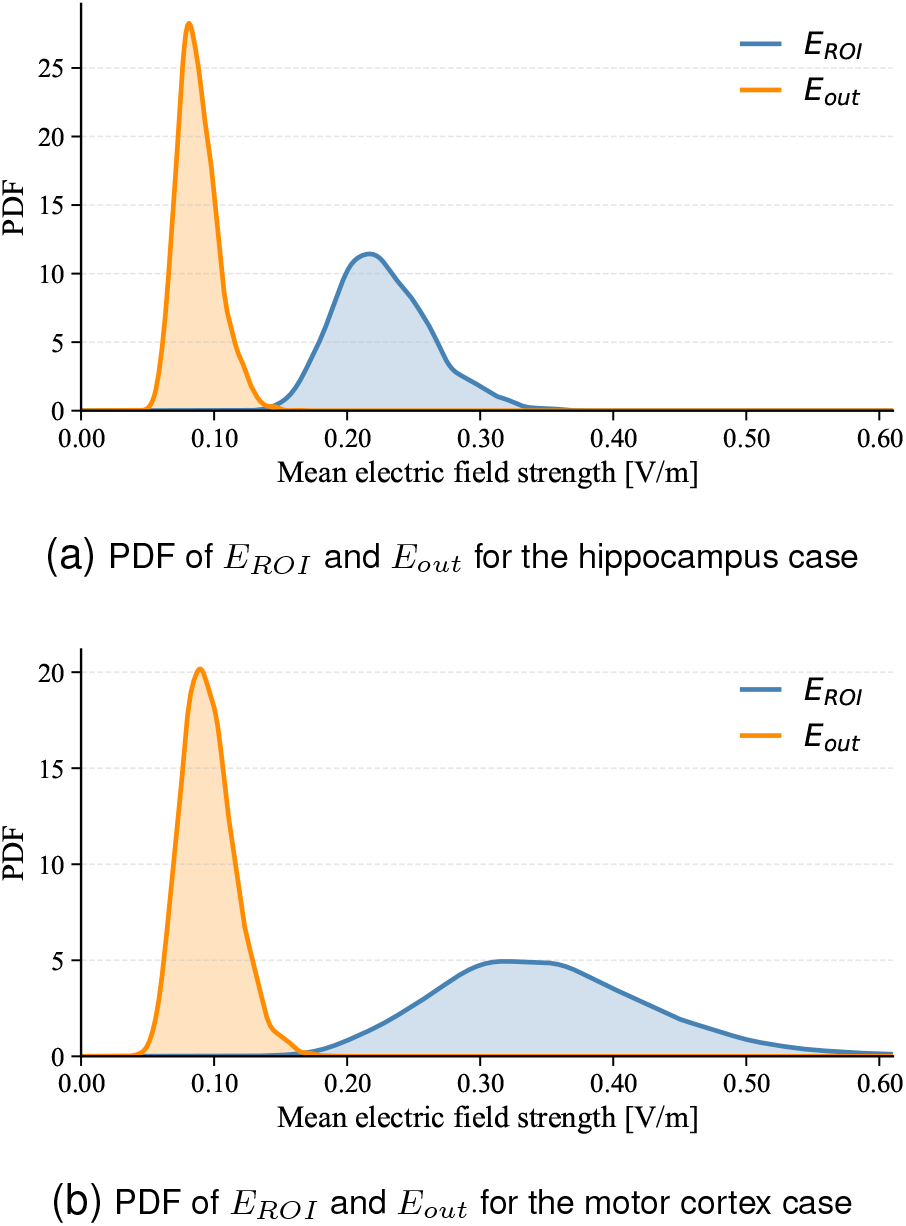
PDFs of ***E***_***ROI***_ and ***E*** _***out***_ for targeted stimulation of (a) the (b) RDM distributions for the motor cortex case hippocampus and (b) the motor cortex. ***E***_***ROI***_ is the mean electric field strength in the target region, whereas ***E***_***out***_ is the mean electric field strength over the midlayer GM outside the target region.

We next assessed the spatial stability of the TI field using the RDM (12), evaluated on the whole midlayer GM Ω_*mid*_ for each of the 4000 samples. The resulting distributions are shown in Fig. 9. For both targets, the RDM values remain below approximately 0.20, with the distribution concentrated around 0.10. RDM values in this range indicate that the spatial pattern of the TI field on the cortical sheet stays consistent with the reference field distribution.

**Fig 9.**
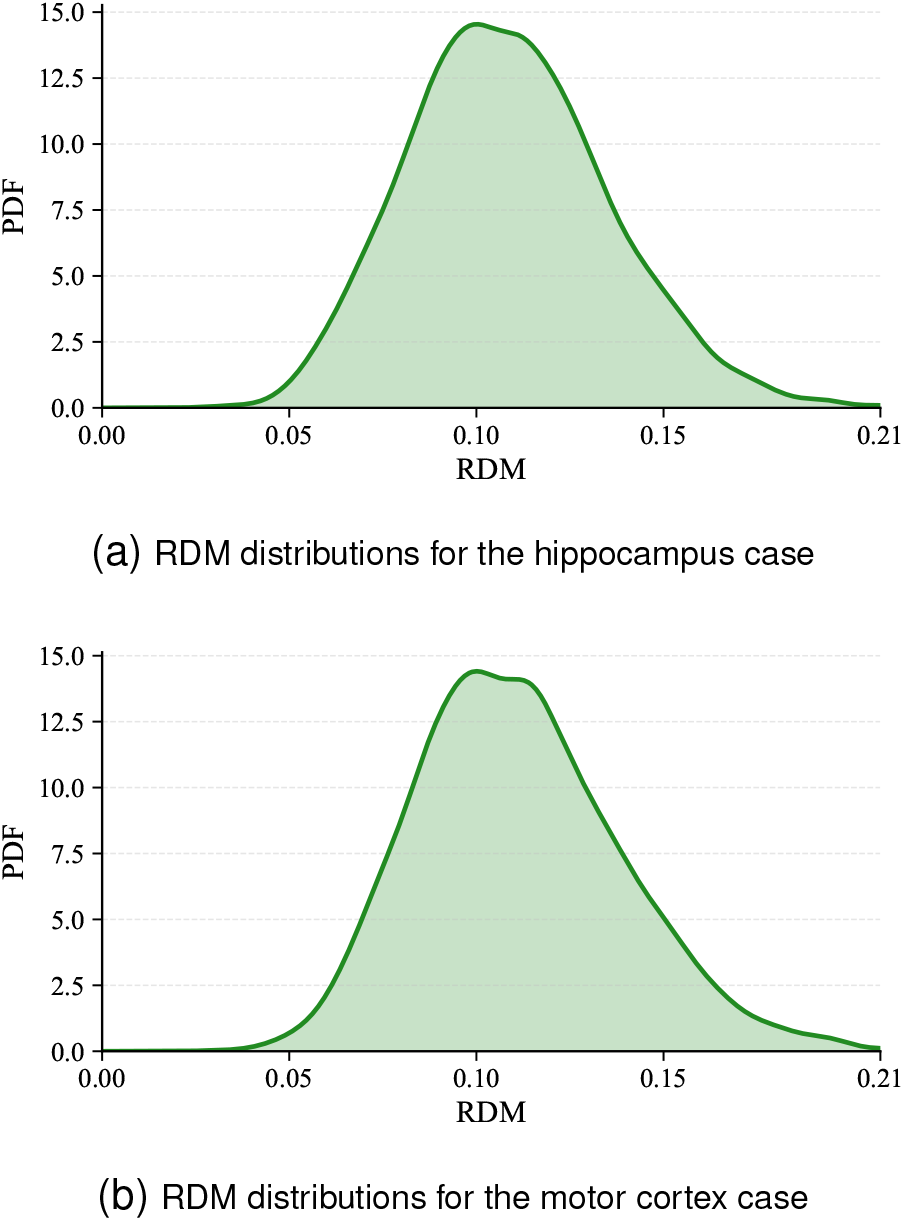
PDFs of the RDM evaluated over the whole midlayer GM **Ω**_***mid***_ for the hippocampus and motor cortex targeted stimulation.

### E. Generalization across Head Models

To assess whether the main findings depend on a single anatomy, we additionally included three individualized head models derived from subjects from the Human Connectome Project dataset [48]. The three additional head models were selected to span the variability of head size, corresponding to maximal (HCP-max), median (HCP-med), and minimal (HCP-min). Each model was processed using the same modeling pipeline as the reference case, and for each head model and each target, the electrode montage was independently optimized using the leadfield-free method proposed in [19] under the same settings and uncertainty definitions as in Section II-A.

Table III reports the contributions of the three uncertainty categories and the residual interaction terms to the total variance of the TI field. The qualitative hierarchy CU > EP > CE is preserved without exception across the four head models and both targets. CU contributes between 88.07% and 99.15% of the total variance, EP between 0.41% and 9.93%, and CE never exceeds 2.6% in any case. CU, therefore, remains the dominant source of variability in every case.

**TABLE III.**
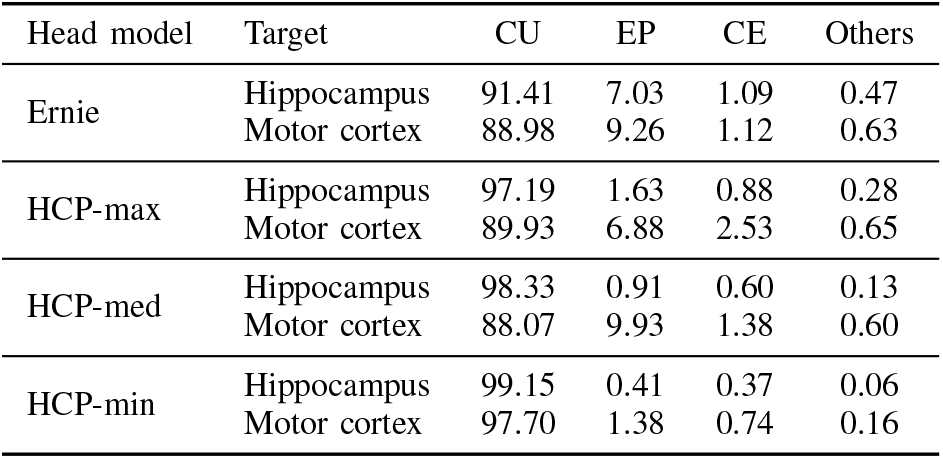
Relative contribution of uncertainty categories (%) across four head models.

| Head model | Target | CU | EP | CE | Others |
| --- | --- | --- | --- | --- | --- |
| Ernie | Hippocampus | 91.41 | 7.03 | 1.09 | 0.47 |
|  | Motor cortex | 88.98 | 9.26 | 1.12 | 0.63 |
| HCP-max | Hippocampus | 97.19 | 1.63 | 0.88 | 0.28 |
|  | Motor cortex | 89.93 | 6.88 | 2.53 | 0.65 |
| HCP-med | Hippocampus | 98.33 | 0.91 | 0.60 | 0.13 |
|  | Motor cortex | 88.07 | 9.93 | 1.38 | 0.60 |
| HCP-min | Hippocampus | 99.15 | 0.41 | 0.37 | 0.06 |
|  | Motor cortex | 97.70 | 1.38 | 0.74 | 0.16 |

We further examined the asymmetry within EP reported in Section III-C across head models. Table IV reports the contribution of each of the four electrodes and of interelectrode interactions to the EP variance for all head models. In every case, a single electrode dominates the EP variance, with the dominant electrode contributing from 66.77% to 78.56%, while the other three electrodes together contribute substantially less. Inter-electrode interactions consistently account for approximately 23% to 25% of the EP variance across cases. The identity of the dominant electrode varies across head models and targets, and the contribution ranking does not follow a consistent ordering of the distance between electrode and target. This indicates that the dominance is not simply determined by geometric proximity to the target.

**TABLE IV.**
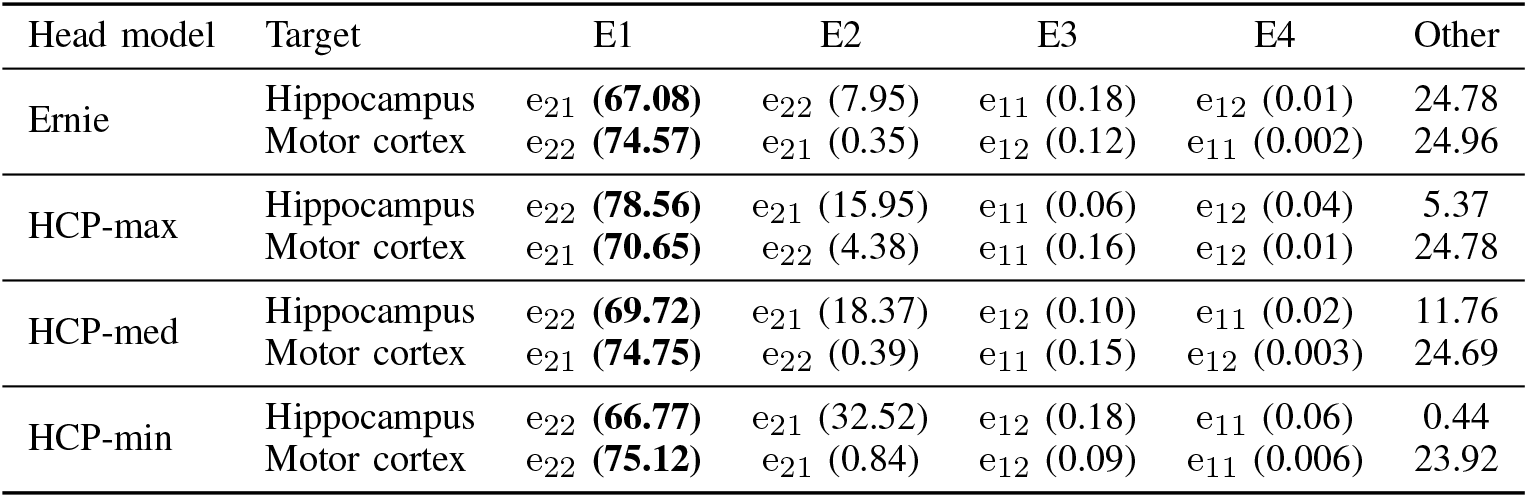
Ranked relative contribution of electrodes across four head models. e1–e4 denote the four named electrodes ranked in descending order of contribution,”other” denotes the combined residual contribution and was not included in the electrode ranking.

## IV. Discussion

This work provides a quantitative account of how different sources of uncertainty shape TIS electric field predictions. Across two representative targets (left hippocampus and motor cortex), the TI field remains focalized over the ROI and comparatively stable across stimulation targets, whereas absolute uncertainty is concentrated in regions of high intensity. This pattern indicates that uncertainty primarily perturbs magnitude rather than spatial pointing, consistent with the mean, standard deviation, and relative standard deviation maps.

This interpretation is further substantiated by the RDM analysis. Across 4000 stochastic realizations and for both targets, the RDM values on the midlayer GM remain below approximately 0.20, with the distribution mass concentrated around 0.10. Such low values indicate that the spatial pattern of the TI field stays close to the reference even when its overall magnitude fluctuates. In other words, uncertainty primarily rescales the TI field rather than relocating its focal point. This spatial robustness measure complements the magnitude evidence from the mean electric field strength inside and outside the target region.

Variance decomposition reveals a clear hierarchy of contributors: CU dominates the field variance, EP has a modest effect, and CE is essentially negligible within the considered ranges. This dominance of CU holds for both deep and superficial targets. For EP, a single electrode accounts for approximately 75% of the variance in the motor cortex case and about 67% in the hippocampus case. This dominant electrode in each case, therefore, constitutes the principal source of variability and should be prioritized for precise localization and stabilization during setup and monitoring. At the tissue level, gray matter is the major contributor to field variance (about 47% for the motor cortex case and about 66% for the hippocampus case), with scalp and compact bone also playing non-negligible roles. These results indicate that more accurate knowledge of tissue conductivities, and of their putative interindividual variations, particularly for gray matter, but also for scalp and compact bone, would offer the largest potential gains in reliability. In comparison, further reductions in session-to-session electrode placement or coregistration errors within the already plausible ranges are likely to yield smaller improvements.

The analyses across four individualized head models support the generalization of the main findings of this work: the variance hierarchy CU>EP>CE and the dominance of a single electrode within EP are preserved in all eight cases considered. The absolute magnitude of each contribution is anatomy-dependent, but the qualitative ranking and the structural mechanisms behind it are preserved. Comparing the deep and the superficial target across the four head models, the geometrical uncertainties contribute systematically more in the superficial case, while CU dominates in both cases.

A practical choice in our sensitivity analysis is to report unnormalized contributions for the spatial Sobol maps. Because local field variance varies by orders of magnitude across space, normalizing at each location can inflate apparent importance where the variance is tiny and suppress it where the variance is large, potentially misrepresenting practical impact. This rationale is noted in our methods and figure caption and is consistent with cases where unnormalized indices are preferable.

From an experimental design standpoint, these findings suggest concrete priorities. First, invest in a better characterization of tissue electric conductivities, particularly for gray matter, scalp, and compact bone, since CU dominates variance and can be highly asymmetric across tissues. Second, improve electrode localization and stabilization (e.g., rigid cap designs, adhesive and strap solutions, in-session drift monitoring). Third, when optimizing electrode setups, explicitly budget for CU in the design objective to obtain robust solutions rather than nominal optima that may be fragile under realistic variability.

Furthermore, we explicitly modeled advanced coregistration and electrode-positioning procedures relying on the availability of individual MRI information and a neuronavigation system. The small contributions of CE and EP within our uncertainty ranges further support the effectiveness of current neuroimaging–based coregistration and electrode-placement technologies. Assessing the impact of discrete electrode positions in standard layouts (e.g., EEG 10-10) and non-linear deformations of flexible EEG caps would be interesting to learn how much less accurate these clinically more scalable solutions would be.

Several limitations should be acknowledged. The stochastic inputs were modeled as independent and bounded according to the literature; the correlated conductivity distributions derived from measurements could be adopted in the future. In addition, the four head models analyzed here are derived from healthy subjects and were selected to span the variability of head size. Patients with altered brain anatomy, such as those with Alzheimer’s disease, are not considered and represent an important direction for future work. Moreover, while the variance decomposition robustly establishes that a single electrode dominates the EP variance, the dominant electrode was not always the closest electrode to the target. The complex interaction between the two electric fields to determine their envelope makes it complicated. Therefore, the variance decomposition does not by itself reveal the full mechanistic origin of this asymmetry. A dedicated analysis of this effect is left for future work.

A related biophysical modeling choice in our framework is the use of isotropic scalar values for tissue conductivities. While currents targeting deep structures like the hippocampus traverse significant WM tracts, our variance decomposition reveals that WM conductivity uncertainty contributes only marginally to the total field variance in the target regions located in GM. Furthermore, according to [49], the conductivity anisotropy only yields small to moderate effects on the simulated electric field.

Recent works have begun to characterize the directional properties of the TI field [50], [51]. In this work, we focus on the scalar amplitude of the maximal modulation envelope, which follows the standard approach for evaluating TIS. The RDM analysis further shows that the spatial pattern of the TI field remains highly consistent under uncertainty, providing indirect support that the underlying spatial structure of the field is preserved, although a direct assessment of orientation stability remains necessary. A dedicated quantitative assessment of orientation stability represents an important direction for future work.

The present analysis is purely computational and builds on SimNIBS as the deterministic forward solver. Experimental validation of the current study would require phantom measurements with head-shaped models and tissue-mimicking materials, in which electrode placement and effective conductivities can be varied in a controlled manner. Establishing such an experimental validation protocol is very challenging and part of future work.

Despite these limitations, the results are encouraging for TIS application in human subjects: within realistic uncertainties, the TIS envelope stays focused and selective, and spatial targeting is robust. The analysis highlights that improving knowledge and personalization of tissue conductivity values is likely to yield the greatest gains in prediction accuracy and stimulation reliability.

## V. Conclusion

In summary, an adaptive PCE framework enabled efficient, quantitative uncertainty and sensitivity analysis for TIS. For both hippocampus and motor-cortex targets, we observed focal mean fields and low leakage, with uncertainty concentrated in high-intensity regions. Total variance is driven primarily by tissue-conductivity variability, whereas electrode placement and coregistration error contribute relatively small amounts in the studied ranges. The quantitative metrics confirm strong dose selectivity and stable spatial targeting under realistic uncertainty.

These findings support a pragmatic recommendation: prioritize resources on improving the measurement of tissue conductivities, so that achieved dose and focality can be interpreted with known bounds. Furthermore, stable electrode positioning is also helpful for stable TIS stimulation.

Demonstrated on two stimulation targets across four individualized head models spanning a range of head sizes, the qualitative findings of this study are systematic and expected to generalize. Quantitative predictions of the TI field, however, remain anatomy-dependent and should be assessed on a subject-specific basis.

## Data Availability

All data produced in the present study are available upon reasonable request to the authors

